# Pharmacogenetics of efavirenz exposure in cervicovaginal fluid during pregnancy and postpartum

**DOI:** 10.1101/2023.10.15.23297046

**Authors:** Oluwasegun Eniayewu, Uche Azuka, Jonah Ogah, Ebunoluwa Adejuyigbe, Oluseye Bolaji, Adeniyi Olagunju

**Affiliations:** Department of Pharmaceutical and Medicinal Chemistry, Faculty of Pharmaceutical Sciences, University of Ilorin, Ilorin, Nigeria; Federal Medical Centre, Makurdi, Nigeria; Department of Paediatrics and Child Health, Faculty of Clinical Sciences, Obafemi Awolowo University, Ile-Ife, Nigeria; Department of Pharmaceutical Chemistry, Faculty of Pharmacy, Obafemi Awolowo University, Ile-Ife, Nigeria; Department of Pharmacology and Therapeutics, University of Liverpool, Liverpool, United Kingdom

**Keywords:** Cervicovaginal fluid, Pregnancy, Postpartum, Efavirenz, Pharmacogenetics

## Abstract

**Objectives:** Adequate antiretroviral drug distribution into the female genital tract (FGT) could play an important role in reducing the risk of heterosexual and mother-to-child transmission of HIV. In this study, we investigated the combined influence of pregnancy and genetic polymorphisms on efavirenz pharmacokinetics in cervicovaginal fluid (CVF) of women receiving antiretroviral therapy.

**Methods:** A total of 159 women (147 pregnant and 12 postpartum) living with HIV and receiving efavirenz-containing antiretroviral therapy were recruited across two sites in Nigeria (Federal Medical Centre, and Bishop Murray Medical Centre, Makurdi) between 2017-2020. In stage 1, sparse CVF and dried blood spot (DBS) samples were obtained from each participant during pregnancy to assess possible association between drug concentration and *CYP2B6* polymorphisms (516G>T and 983 T>C). In the second stage, participants were stratified into three genotype groups (extensive, intermediate and low metabolisers) and re-enrolled for intensive pharmacokinetic sampling.

**Results:** In stage 1 (88 CVF, 81 plasma and 73 paired samples), *CYP2B6 516G>T* was independently associated with both CVF (β = 997 ng/mL (90% CI: 598, 1357), *p =* 5.7 x 10^-5^) and plasma (β = 1400 ng/mL (90% CI: 1051, 1748), *p =* 5.7 x 10^-9^) efavirenz concentration during pregnancy. In the second stage (12 pregnant, 12 postpartum), median (IQR) efavirenz C_min_ in CVF during pregnancy versus postpartum was 243 ng/ml (168-402) vs 447 ng/ml (159-974), C_max_ was 1031 ng/ml (595-1771) vs 1618 ng/ml (675-2695), and AUC_0-24_ was 16465 ng.h/ml (9356-30417) vs 30715 ng.h/ml (10980-43714). Overall, median CVF-to-plasma AUC ratio was 0.34 during pregnancy and 0.46 postpartum. When patients were stratified using *CYP2B6 516G>T*, efavirenz median clearance increased by 57.9% during pregnancy compared with postpartum control (*p* = 0.232) in patients with the *CYP2B6* 516GT genotype. The AUC_0-24h_, C_max_ and C_min_ reduced by 33.8% ((p=0.182), 8.6% (0.175) and 59.5% (0.171) during pregnancy, with values of 20671 ng.h/ml (15993-28712), 1550 ng/ml (1090-2090) and 330 ng/ml (250-440), respectively, compared with 31229 ng.h/ml (27660-41873), 1695 ng/ml (1540-3003) and 814 ng/ml (486-981) during postpartum in this genotype.

Median efavirenz C_min_ in CVF was 1.93 and 3.55 times higher than the PBIC_90_ of 126 ng/ml in the pregnant and postpartum cohorts, respectively.

**Conclusions:** Efavirenz is well distributed into the CVF, and both pregnancy and polymorphisms in its disposition genes affect CVF exposure.

## Introduction

The understanding of antiretroviral pharmacokinetics in the female genital tract (FGT) is of growing interest in the HIV prevention field because of its importance in designing more effective therapeutic options to reduce serodiscordant and mother-to-child HIV transmission[1]. Although a number of clinical studies that focus on quantifying antiretroviral drugs in the FGT have been reported in the literature[2–6]. ARV characterization in this compartment has not yet been well understood. While pregnancy and polymorphisms have been established to alter ARV plasma exposure[7–10], the knowledge of the effect of these factors on ARV disposition in the FGT is very limited. With evidence of an ongoing risk of viral shedding in FGT, even in patients on suppressive antiretroviral therapy (ART) despite undetectable plasma viremia[11–13], there is a need to further understand antiretroviral dynamics in this compartment to enable the design of effective therapies.

MTCT HIV-1 transmission is essentially minimized by suppressing HIV replication and reducing plasma viral load to an undetectable level, particularly at the early stages of pregnancy[14]. Vaginal viral shedding in the instance of suppressed plasma vireamia and in patients newly placed on ART drugs can potentially increase the risk of heterosexual and MTCT HIV transmission[15,16]. As of 2022, an approximated 76.4% (29.8 million of the 39 million people living with HIV globally) were reported to be receiving antiretroviral therapies (ART), including at least 8 out of every 10 pregnant women with HIV [17]. Despite this high access to ART, new HIV transmission among children remains substantial, most of it through the perinatal route. Since a substantial proportion of MTCT occurs at the time of delivery via contact with infected female genital fluids [18,19], a significant percentage of these new pediatric HIV infections can be prevented by having a better understanding of ARV disposition in the FGT and factors that could affect FGT drug exposure.

The importance of improving the current understanding of ARV pharmacokinetics within and outside the context of pregnancy and pharmacogenetics cannot be overemphasized in the context of designing more effective HIV preventive therapy. However, obtaining robust pharmacokinetic data to achieve this has been constrained by the invasiveness and increased cost associated with sampling genital tissues[20] and the limitations of standardizing existing bioanalytic methods used to quantify ARVs in these compartments[21]. To circumvent these challenges,, cervicovaginal fluids (CVF are used as an acceptable surrogate for vaginal tissue concentrations of ARVs[20]. In our laboratory, we recently validated a minimally invasive LC-MS/MS method to quantify the HIV drug efavirenz in cervicovaginal fluid collected on flock swabs[6], which improved our ability to conduct intensive clinical pharmacokinetic studies with CVF.

In this component of the VADICT study, whose protocol was previously published [22], we set out to characterize efavirenz disposition in the FGT and determine the impact of the combined effect of pregnancy and CYP 2B6 Single Nucleotide Polymorphisms (SNPs) on the FGT disposition of the drug.

## Methods

### Study sites, participants, and ethics

Participants in the VADICT study were recruited from four sites in Benue State, Nigeria (Federal Medical Centre, Makurdi; Bishop Murray Medical Centre, Makurdi; St. Monica’s Hospital, Adikpo; and St. Thomas’ Hospital, Ihugh) between December 2017 and September 2020. Pregnant or recently postpartum women living with HIV who were at least 18 years old, receiving WHO recommended first-line ART regimen, not taking other medications with known or suspected interaction with ART components, and who had no severe illness were eligible. The study was approved by the National Health Research Ethics Committee, Abuja, Nigeria (approval number: NHREC/01/01/2007-05/06/2017). The study is registered on ClinicalTrials.gov (NCT03284645) and the full protocol has been published elsewhere [22]. Participants for parts of the study reported here were recruited only from Makurdi sites where ultra-low temperature facilities were available.

### Study design

The VADICT study was an observational cohort study with a pharmacokinetic component conducted in two stages. In the first stage, we assessed association between established functional single nucleotide polymorphisms (SNPs) in efavirenz disposition gene *CYP2B6* (516G>T and 983 T>C) and mid-dose efavirenz concentration in both cervicovaginal fluid and plasma during pregnancy. For stage two, participants were stratified into three metabolizer groups using the independently associated SNP based on the number of variant alleles present: ‘slow’ (two), ‘intermediate’ (one), and ‘fast’ (none) . Participants from each group who still satisfied the eligibility criteria at the time the pharmacogenetic data became available were invited for re-enrollment into the intensive pharmacokinetic stage.

### Sample collection and handling

Paired plasma and cervicovaginal fluid samples were collected from each participant at a single recorded timepoint after the last dose. About 1-2mL of plasma was obtained from each participant by centrifugation (4000 rpm) of venous blood samples and transferred into cryovials. Cervicovaginal fluid samples were collected with flocked swabs by a female nurse as described in our paper on this method [23]. In summary, participants laid down on their backs for 5 min before each sample collection to allow pooling of fluid in the back of the vagina. The swab was gently inserted approximately 3 inches into the vagina at first, and inserted further until it touched the back of the posterior fornix. The swab was gently rubbed against the mid-vaginal walls for at least 30 sec, withdrawn and immediately transferred into a cryovial.

Participants who were re-enrolled in stage 2 after stratification into slow, intermediate and fast metaboliser underwent intensive pharmacokinetic sampling. CVF and dried blood spot (DBS) samples were collected at six different time points over a single dosing period (pre-dose and at 0.5, 1, 2, 4, 8, and 12 h after dose) following an observed evening dose of ART regimen containing 600 mg efavirenz. Within a few minutes of CVF collection, DBS samples were prepared by spotting drops of whole blood collected on Whatman protein saver cards by finger prick using a 2 mm safety lancet (BD, Oxford, Oxfordshire, UK). DBS cards were dried at ambient temperature (18-25°C) for at least 2 hours, and thereafter packed in ziplock bags with desiccants. Standard operating procedures were prepared for all sample collection processes, and these were performed by trained staff involved in routine care of study participants at each site. All samples were stored at -80°C at the Federal; Medical Centre, Makurdi until transferred to the Translational Pharmacokinetics Research Laboratory, Obafemi Awolowo University, Ile-Ife using Arctic Express® Dry Shipper (Thermo Scientific, Waltham, MA, USA) where they were stored at -80 °C until analysis.

### Pharmacogenetic analysis and drug quantification

Two *CYP2B6* SNPs (*CYP2B6* 516G > T and CYP*2B6* 983 T > C) known to be associated with efavirenz plasma exposure were included in this analysis. Genomic DNA was extracted from DBS using the QIAamp DNA Blood Mini Kit (Qiagen, Germantown, USA) and quantified using Nanodrop® spectrophotometer (Thermo Fisher Scientific Inc., Wilmington, DE, USA) and stored at -70°C. Genotyping was performed by real-time PCR based allelic discrimination using TaqMan® SNP genotyping assays for *CYP2B6* 516G>T (rs3745274 with product ID C_7817765_60) and *CYP2B6* 983T>C (rs28399499 with product ID C_60732328_20) on a DNA Engine Chromo4 system (Bio-Rad Laboratories, Inc., Hercules, CA, USA). The three-step PCR protocol included a 15-minute denaturation step at 95°C, 50 cycles of amplification at 95°C for 15 s each, and a 1-minute annealing step at 60°C. Opticon Monitor® version 3.1 (Bio-Rad Laboratories, Inc., Hercules, CA, USA) enabled genotype calls based on allelic discrimination plots.

Determination of efavirenz concentration in CVF, plasma and DBS samples was conducted on a TSQ Quantum Access LC-MS/MS system (Thermo Fisher Scientific Inc., Hemel Hempstead, Hertfordshire, UK) at the Bioanalytical Laboratory, Faculty of Pharmacy, Obafemi Awolowo University, Ile-Ife, Nigeria using previously reported methods [24,23]. To estimate plasma efavirenz concentration from DBS concentration, the equation originally reported by Eyles et al ([25]) which accounts for the effect of fraction unbound to plasma proteins and haematocrit was used.

### Statistical analysis

Compliance with Hardy–Weinberg Equilibrium was tested as previously described. Efavirenz plasma and CVF concentration dataset were subjected to Kolmogorov-Smirnov normality test prior to statistical analysis. Spearman correlation was performed to test the relationship between continuous variables. Variables associated with efavirenz concentration in plasma and CVF were determined using the univariate linear regression analysis. Variables that showed significant association with efavirenz plasma and CVF concentration in the univariate linear regression analysis were included in a multivariate stepwise linear regression analysis. Differences in efavirenz plasma and CVF concentrations and pharmacokinetic parameters between patient groups were investigated using 1-way analysis of variance (ANOVA) and Mann–Whitney *U* test All these analyses were performed using IBM SPSS Statistics version 29.0.1.0 (171) (IBM, Armonk, New York) and GraphPad Prism 10 (GraphPad Software, Inc., La Jolla, California).

## PResults

### Study participants

A total of 147 pregnant women contributed sparse pharmacokinetic samples in stage 1 at a mean (SD) gestational age of 34.5 weeks (3.12), age 28.9 years (4.9) and body weight 64.3 kg (11.9). Median (range) duration on therapy prior to enrolment into the preliminary phase of the study was 40.9 months (0.03 - 149). Twelve pregnant women aged 28.9 years (4.9) old with body weight 65.7 kg (7.00) participated in the intensive pharmacokinetic sampling of stage 2 at gestational age of 26.0 weeks (6.70). Postpartum intensive pharmacokinetic sampling was conducted in 12 women aged 32.9 years (5.1) at 37.1 weeks (4.1) after delivery. Total duration on ART prior to enrolment for stage 2 was 41.2 weeks (36.7) during pregnancy and 55.8 weeks (27.3) for postpartum women. All study participants received efavirenz (600 mg), lamivudine (300 mg) and tenofovir disoproxil fumarate (300 mg) fixed dose combination ART.

A total of 169 sparse samples (88 CVF, 81 plasma, and 73 paired samples at third trimester), 168 intensive CVF samples (84 samples each from the 12 pregnant and 12 postpartum women), and 98 DBS (48 samples each from 7 pregnant and 7 postpartum women) were available for analysis. The *CYP2B6* 516G>T SNP was in Hardy-Weinberg equilibrium within the study population.

### Are *CYP2B6* 516G>T and 983 T>C associated with efavirenz CVF concentration?

In stage 1, 88 sparse CVF and 81 plasma samples were obtained from a total of 92 participants during pregnancy (73 paired samples) to assess possible association between efavirenz concentration and *CYP2B6* polymorphisms (516G>T and 983 T>C). CVF and plasma samples were collected 14.45 h (1.30-17.02) after the last dose. .The median (IQR) CVF and plasma efavirenz concentrations were 788 ng/mL (502-1542) and 1487 ng/mL (1041-2363), respectively. There was significant correlation in efavirenz CVF and plasma concentration at 90% confidence interval (*p*= 1.32 x 10^-6^, Pearson’s r = 0.51 (0.37-0.66). The average CVF-to-plasma ratio computed from the 73 paired sparse samples was 0.55.

The result indicates that only *CYP2B6 516G>T* is independently associated with efavirenz CVF and plasma concentration during pregnancy. The unstandardized regression coefficient (β)(90% confidence internal, *p*-value) for this association is 997 ng/mL (598-1357, *p* = 5.7 x 10^-5^) and 1400 ng/mL (1051-1748, *p* = 5.7 x 10^-9^), respectively. Notably, *CYP2B6 516G>T* alone accounted for 20 % (adjusted R square = 0.20) and 40 % (adjusted R square = 0.40) of the observed differences in the efavirenz CVF and plasma concentrations during pregnancy, respectively. The association between *CYP2B6 516G>T* and efavirenz CVF as well as plasma concentration is presented in Figure 1.

**Figure 1:**
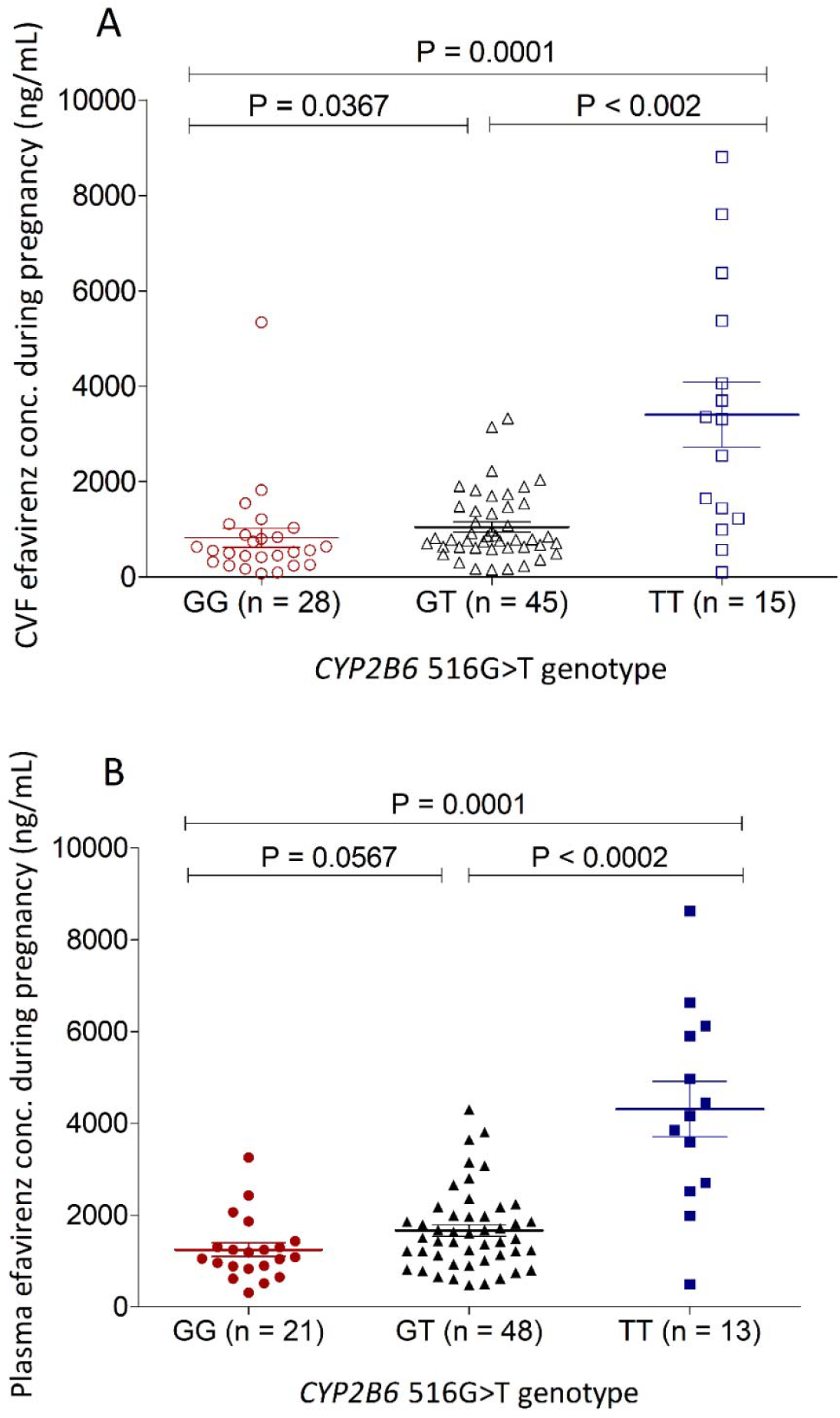
Association of pooled *CYP2B6* 516G>T with EFV Cervicovaginal fluid (CVF) concentrations during pregnancy. *P* values on figures are for Mann-Whitney U / Kruskal Wallis test. GG; patients with two normal function *CYP2B6* allele, GT; patients with one normal and one decreased function *CYP2B6* allele, TT; patients with two decreased function *CYP2B6* alleles

No significant association was observed between efavirenz CVF concentration and *CYP2B6 983T>C* (*p* = 0.573), age (*p* = 0.134), body weight (*p* = 0.716), body mass index (*p* = 0.873), and time after dose (*p* = 0.729). Similarly, *CYP2B6 983T>C*, age, body weight, body mass index, and time after dose were not significantly associated with efavirenz plasma concentration.

Upon stratification based on *CYP2B6 516G>T*, we observed significant differences (*p* < 0.0001) in efavirenz median (IQR) CVF concentrations: GG (n=28), 559 ng/mL (342-874); GT (n= 45), 791 ng/mL (630-1475); TT (n=15), 3310 ng/mL (1332-4718). Similarly, plasma concentrations varied based on *CYP2B6 516G>T* (*p* < 0.0001): GG (n= 21), 1085 ng/mL (888-1305); GT (n= 48), 1495 ng/mL (1095-1983); TT (n=13), 4164 ng/mL (2698-5903). Only *CYP2B6 516G>T*, was used to stratify study participants invited for the intensive pharmacokinetic phase.

### What is the combined influence of pregnancy and genetics on efavirenz CVF exposure?

In the second stage, 12 pregnancy women (6GG and 6GT) and 12 postpartum women (4GG, 5GT and 3TT) were invited for intensive pharmacokinetic sampling. Efavirenz concentration-time profile during pregnancy and postpartum in the combine population and in *CYP2B6* 516GG *and CYP2B6* 516GT groups are presented in Figure 2A. The corresponding pharmacokinetic parameters are presented in Table 2. In pooled analysis, we observed a 87.7% increase (*p*-value at 90% confidence interval = 0.072) in the median clearance, 46.4 % decrease in AUC_0-24h_ (*p* = 0.042), 36.3% decrease in C_max_ (*p* = 0.069), and 45.6 % decrease in C_min_ (*p* = 0.085). However, upon stratification based on *CYP2B6 516G>T*, efavirenz median clearance increased by 57.9% during pregnancy compared with postpartum control (*p* = 0.232) in patients with the *CYP2B6* 516GT genotype. The AUC_0-24h_, C_max_ and C_min_ reduced by 33.8% ((p=0.182), 8.54% (0.175) and 59.9% (0.171) during pregnancy, with values of 20671 ng.h/ml (15993-28711), 1551 ng/ml (1087-2086) and 330 ng/ml (251-442), respectively, compared with 31229 ng.h/ml (27660-41873), 1695 ng/ml (1540-3003) and 814 ng/ml (486-981) during postpartum. In the contrary, efavirenz CVF exposure was marginally increased during pregnancy in patients with the *CYP2B6* 516GG genotype (Table 2). We could not compare between the pharmacokinetic parameters of patients with the *CYP2B6* 516TT genotype in the pregnancy and postpartum cohorts because there were no patients with the *CYP2B6* 516TT genotype in the pregnancy cohort.

**Table 1:**
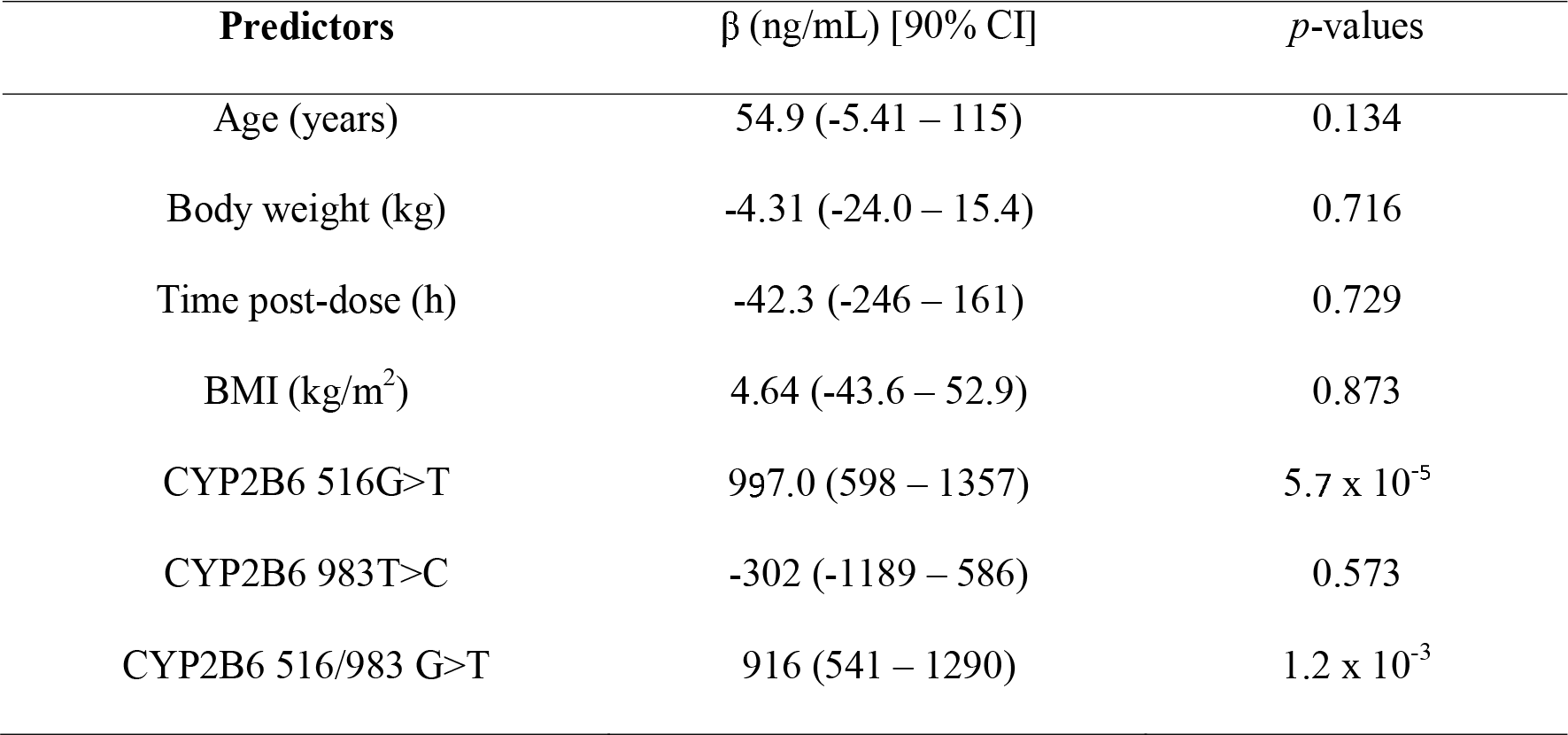
Univariate linear regression analysis of factors influencing efavirenz CVF disposition in pregnant women.

**Table 2:** Efavirenz CVF pharmacokinetics in genetically stratified pregnant and postpartum women Results are presented in median (Interquartile range, IQR).

| PK parameters | Combined (n=12) | CYP2B6 516GG (n = 6) | CYP2B6 516GT (n = 6) |  |
| --- | --- | --- | --- | --- |
| **Pregnancy** |  |  |  |  |
| C<sub>min</sub> (µg/mL) | 243 (168 – 402) | 169 (164 – 242) | 330 (250 – 440) |  |
| C<sub>max</sub> (µg/mL) | 1031 (595 – 1771) | 664(596 – 1069) | 1550 (1090 – 2090) |  |
| AUC<sub>0-24h</sub> (µg.h/mL) | 16465 (9356 – 30417) | 11577 (7507 – 13284) | 20671 (15993 – 28712) |  |
| CL/F (L/h) | 36.6 (19.7 – 65.1) | 52.0 (42.0 – 79.9) | 30 (21 – 37.6) |  |
| T<sub>max</sub> (h) | 8.00 (7.00 – 9.00) | 8.00 (4.00 – 12.0) | 8.00 (8.00 – 8.00) |  |
| **Postpartum** |  |  |  |  |
|  | Combined (n=12) | CYP2B6 516GG (n = 4) | CYP2B6 516GT (n = 5) | CYP2B6 516TT (n = 3) |
| C<sub>min</sub> (ng/mL) | 447 (159 – 974) | 151 (116 – 225) | 814 (496 – 981) | 972 (587 – 2383) |
| C<sub>max</sub> (ng/mL) | 1618 (675 – 2695) | 636 (583 – 1107) | 1695(1540 – 3003) | 2587 (1633 – 4365) |
| AUC<sub>0-24h</sub> (ng.h/mL) | 30715 (10980 – 43714) | 10630(9848 – 15875) | 31229 (27660 – 41873) | 39673 (25147 – 64938) |
| CL/F (L/h) | 19.5 (13.8 – 54.7) | 57.0 (46.0 – 61.1) | 19.0 (14.0 – 22.0) | 15 (10.9 – 35.8) |
| T<sub>max</sub> (h) | 8.0 (8.0 – 8.0) | 8.0 (7.0 – 8.0) | 8.0 (8.0 – 8.0) | 8.0 (8.0 – 8.0) |

**Figure 2:**
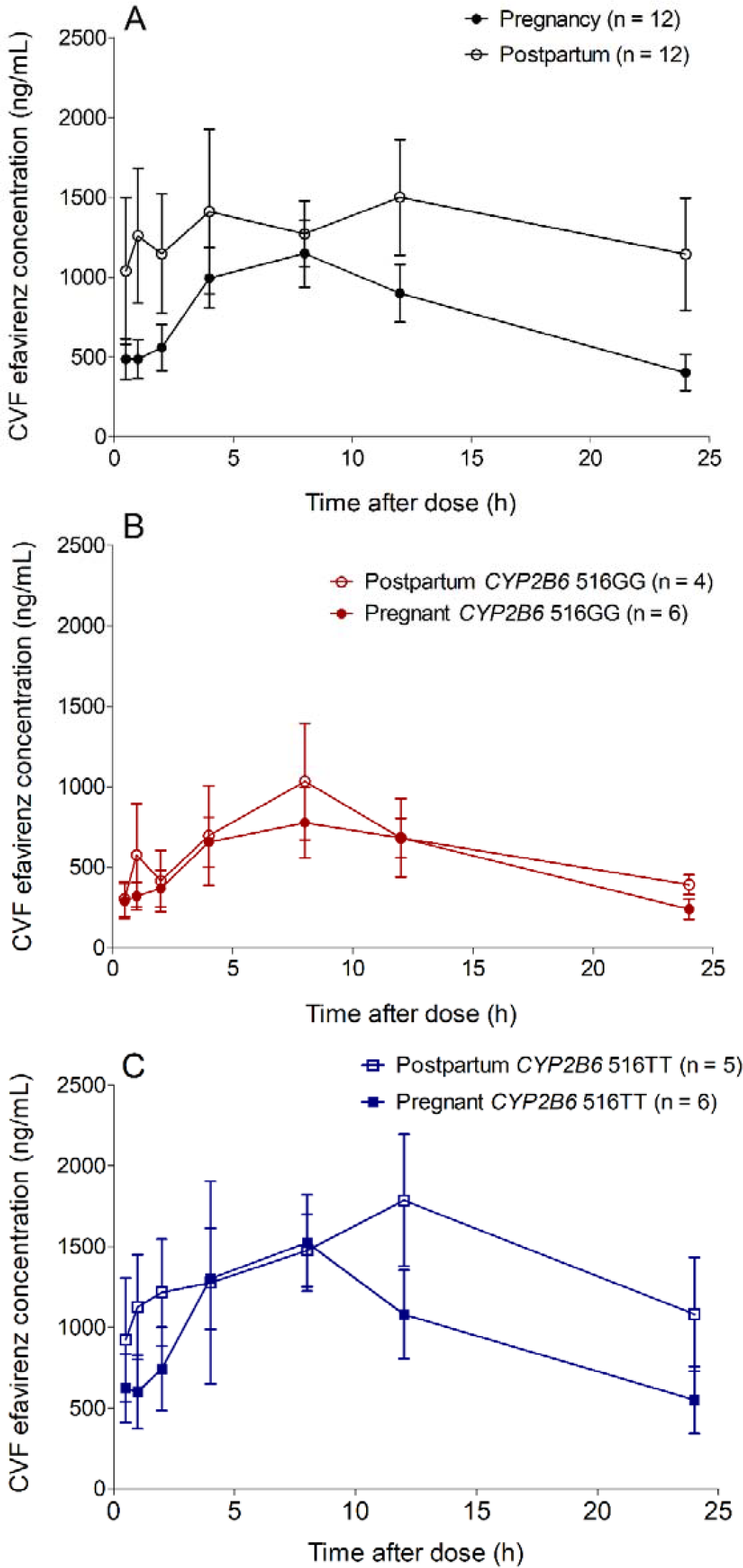
Efavirenz CVF concentration–time profiles (mean, SEM) during pregnancy and postpartum

### Efavirenz CVF and Plasma pharmacokinetics during pregnancy and postpartum

Efavirenz pharmacokinetic parameters differed significantly in CVF and plasma. Median (IQR) pharmacokinetic parameters in CVF (n=12) versus plasma (n=7) were AUC_0-24_, 16465 ng.h/mL (9356-30417) versus 48983 ng.h/mL (41414-54311); C_max_, 1031 ng/mL (585-1771) versus 5080 ng/mL (4535-5364); and C_min_, 243 ng/mL (168-402) versus 844 ng/mL (534-1109) during pregnancy, and 30715 ng.h/mL (10980-43714) versus 67339 ng.h/mL (57492-106868); C_max_, 1618 ng/mL (675-2695) versus 5174 ng/mL (4643-6988); and C_min_, 447 ng/mL (159-974) versus 2192 ng/mL (1637-6485) in postpartum women. CVF-to-plasma C_max_ and AUC_0-24_ ratios were 0.20 and 0.34 during pregnancy, and 0.31 and 0.46 during postpartum. Time to reach efavirenz maximum plasma versus CVF concentration were 2.00 h (2.00-4.00) vs 8.00 h (7.00 – 9.00) and 2.00 h (2.00-2.00) vs 8.00 h (8.00 – 12.0) in pregnant and postpartum women, respectively.

## Discussion

The female genital tract (FGT) is an important sanctuary site where a significant proportion of MTCT occur during delivery. Strategies to reduce HIV transmission rates focuses on reducing the amount of virus in the FGT with antiretroviral therapy [26]. A good knowledge of the factors that affect the disposition of antiretroviral drug into the FGT of HIV-infected pregnant women can potentially aid accurate assessment of antiretroviral-exposure relationships and improve maternal-infant survival outcomes. The impact of pregnancy and host genetics on the disposition of ART into the FGT has not been well studied. In this study, we performed a clinical pharmacokinetic study to describe efavirenz FGT disposition in genetically defined pregnant and postpartum women.

Our findings indicates that efavirenz distributes rapidly into the cervicovaginal fluid (CVF) of pregnant and postpartum women after oral dosing. Although efavirenz minimum effective concentration in the FGT has not been established, concentrations exceeding the efavirenz PBIC_90% (_126 ng/ml) for wild-type HIV-1 measured within 30 minutes post-drug administration in 83% of pregnant and postpartum women studied may suggest that efavirenz administration at 600 mg may be sufficient to reduce HIV MTCT at the time of delivery. Comparatively, we observed that efavirenz CVF disposition is positively correlated with plasma exposure (Pearson’s r = 0.76 (0.65 – 0.85), R^2^ = 0.58, P-value < 0.0001), with CVS:plasma efavirenz AUC_0-24h_ observed to be 0.34 and 0.46 in the pregnant and postpartum women who contributed the intensive pharmacokinetic samples. Some other studies have also reported good penetration of antiretroviral drugs into the FGT. For example, CVF-plasma concentration ratios of 2.3 (1.4 – 4.1), 3.2 (1.2 – 8.5) and 5.2 (1.2 – 22.6) have been reported for raltegravir [27], lamivudine [2] and tenofovir [2], respectively.

Factors such as molecular weight and protein binding can determine the extent of drug distribution into the FGT. Efavirenz is a low molecular drug and it is expected to be able to penetrate the female genital tissues and fluid. Drugs penetration into the female genital tissues appear to be influence by its protein binding ability. Highly protein bound drugs generally tends to have more capacity to penetrate the FGT compared to poorly bound drugs [28]. Efavirenz protein binding in the CVF was not determined in this study. However, based on previous reports, the concentrations of drug binding proteins (mainly albumin and alpha 1-acid glycoproteins) has been previously reported to be approximately 1% of what is obtained in blood plasma [29]. Also, maraviroc CVF protein binding have been reported to be 10% of plasma protein binding [30]. Based on this information, it can be assumed that efavirenz CVF protein binding is limited hence the high concentration of the drug in the CVF.

Our results indicate a 46% reduction in efavirenz CVF AUC_0-24 h_ during pregnancy relative to postpartum values. Both efavirenz CVF C_min_ and C_max_ during pregnancy were substantially lower than postpartum by 45.6% and 36.3% respectively. Efavirenz clearance from the CVF was markedly increased (87.6% increment) during pregnancy compared to postpartum control. Reduced antiretroviral CVF exposure during pregnancy may be associated with sub-optimal suppression of viral replication in the FGT and increased risk of MTCT of HIV, particularly among HIV-positive pregnant women who were diagnosed of HIV and commence ART in late pregnancy. The onset of pregnancy is well known to be associated with maternal physiological changes [31]. These changes could alter drug disposition and may explain the reduction in the CVF exposure of efavirenz observed in this study.

Efavirenz is predominantly metabolized by CYP2B6 [32] and the *CYP2B6* 516G>T genotype has been shown to reduce the rate of efavirenz biotransformation to its major metabolite, efavirenz 8-hydroxylation [33].

We found that on *CYP2B6* 516G>T genotype was strongly associated with efavirenz CVF disposition in pregnant and postpartum women. Efavirenz concentration in the CVF varies depending on *CYP2B6* 516GT genotypes during pregnancy and postpartum, with the lowest CVF exposure measured in subjects stratified as *CYP2B6* 516GG genotype. The overall CVF exposure of efavirenz in pregnant women classified as CYP*2B6* 516GG genotype was lower by 43.9% compared to those classified as CYP*2B6* 516GT genotype. On the other hand, the overall efavirenz CVF exposure in postpartum women classified CYP*2B6* 516TT genotype exceeded those classified as CYP*2B6* 516GG and CYP*2B6* 516GT by 273% and 27.0%, respectively. This findings suggests that *CYP2B6* Single Nucleotide Polymorphisms (SNPs) play a role in efavirenz CVF disposition

Efavirenz CVF C_min_ was above the reported value of 126 ng/mL for the wild type HIV-1 in 75% of postpartum women with the CYP*2B6* 516GG genotype, compared to 100% in pregnant women with the same genotype. The genotype-dependent changes in the efavirenz CVS exposure observed in this study may therefore be explained by possible genotype-dependent differences in efavirenz autoinduction in the human female genital tract. Unfortunately, the expression of CYP2B6 isoenzyme and its potential autoinduction in the human female genital tract or any other genetic factors was not directly assessed in this study. This is considered one of the limitations of this study and further studies in this area are recommended. Our results supports *CYP2B6* 516G>T-guided dosing of efavirenz in pregnant women.

The findings from this study need to be interpreted with caution and in the context of the limitation associated with the conduct of this study. Firstly, the women who contributed the intensive pharmacokinetic samples during pregnancy were different from cohorts who contributed the postpartum samples. Since the cohorts of the pregnant women were not used as their own postpartum control, there may be some bias in the interpretation of the effect of pregnancy and host genetics on efavirenz CVF exposure.

In conclusion, the study established pregnancy and *CYP2B6* SNPs play an important role in efavirenz disposition into the female genitalia.

## Data Availability

All data produced in the present study are available upon reasonable request to the author

## Acknowledgements

The authors acknowledge bioanalytical facility support from the Obafemi Awolowo University Bioanalytical Laboratory, Nigeria and the University of Liverpool, United Kingdom.

## Figure Legends

Figure 1: Association of pooled *CYP2B6 c*. 516/985 with EFV CVS concentrations during pregnancy in the preliminary phase. *P* values on figures are for Mann-Whitney U / Kruskal Wallis test.

Figure 2: Efavirenz CVF concentration–time profiles (mean, SEM) during pregnancy and postpartum (A). Comparison shows decreased exposure during pregnancy (n = 12) compared with postpartum (n = 12). Upon stratification of patients based on CYP2B6 516G>T status showed decreased exposure during pregnancy (n = 6) compared with postpartum (n = 4) in patients with the GG genotype(B). Similarly, efavirenz exposure decreased in pregnant women (n=6) compared to postpartum women (n=5) with the GT genotype (C). None of the pregnant women recruited into the intensive phase of this study had the TT genotype. Median (range) C_min_, C_max_, and AUC_0-24_ of postpartum women (n=3) with the TT genotype status were 972 (587-2383), 2587 (1633-4365) and 39673 (25147-64938), respectively.

